# A 10 Years Update of Effects of Exercise on Depression Disorders – in otherwise healthy adults: A Systematic Review of Meta-Analyses and Neurobiological Mechanisms

**DOI:** 10.1101/2024.08.28.24312666

**Authors:** Henning Budde, Nina Dolz, Anett Mueller-Alcazar, Bruna Velasques, Pedro Ribeiro, Sergio Machado, Mirko Wegner

## Abstract

**Background:** Depression is one of the most common mental illnesses worldwide and is a major burden for those affected. As conventional therapies do not always work and are also associated with side effects, alternative treatment methods are urgently indicated. In the past, exercise has established itself as a seemingly good alternative treatment method. The aim of this work is to provide a state of the art review and to check whether there are new findings since the publication of the article by Wegner et al. [1].

**Methods:** A systematic literature search was conducted in which relevant literature was searched in databases such as PsycINFO, PsychARTICLES, PubMed, CINAHL Complete, SocINDEX, SPORTDiscus and Psyndex. The effect sizes were calculated and the methodological quality was assessed using the AMSTAR-2 criteria. Finally, the neurobiological explanations for the effect of exercise on depression are discussed.

**Results:** Eleven meta-analyses met the inclusion criteria, with the total sample consisting of 16.255 participants and 229 individual studies. The most frequently implemented intervention was aerobic exercise, while the intervention in the control groups was usually no treatment, waiting list, or attention/activity placebo. The pooled results indicate a moderate clinical effect, suggesting the positive effect of exercise and physical activity in reducing depressive symptoms (SMD = -0.61, 95% CI [− 0.78; -0.43], *p* = <0.01).

**Conlcusion:** The consistently positive, moderate effects observed in the present study make exercise and physical activity a promising and supportive alternative for adults with depression. The positive effect of exercise and physical activity could potentially be explained by neurological changes. However, the exact mechanisms underlying the antidepressant effects are still unclear.

## 1 Introduction

The World Health Organization (WHO) not only considers physical inactivity to be the epidemic of the 21st century, it is also the fourth most important risk factor for mortality and is associated with the spread of noncommunicable diseases [2]. According to the World Obesity Federation (WOF), it is estimated that 51% of the world’s population will be overweighted by 2035 [3]. According to BRFSS reports, only 48.8% of adults in the US reach the minimum level of physical activity (PA) required for good health. This places a major burden on the healthcare system [4]. The relationship between PA as well as exercise and health outcomes is widely known [5]. Measures to promote PA have contributed significantly to the alleviation of physical complaints. Similar effects can be observed for mental health: People who exercise regularly suffer less from anxiety, fatigue, and cognitive impairment [1, 4, 6]. Although mental illnesses such as depression have the potential to significantly impair quality of life depressive disorders have become a widespread health problem worldwide [7, 8]. The statistics show that between 2015 and 2019, the prevalence of depression increased significantly without a corresponding increase in the number of treatments. Although more and more time is passing, the problem is getting worse [9]. The treatment of depression, whether pharmacological or cognitive behavioral therapy, has various problems. One problem about pharmaco treatment is the concern about drug-related side effects (such as constipation, diarrhea, dizziness, headaches, insomnia, nausea, decreased sexual desire and drowsiness) or, more generally, the social stigma associated with mental disorders [1, 10, 11]. These can significantly impair the acceptance of treatment and adherence to therapy [11]. Although there are many effective antidepressants, over 50% of patients do not respond to the first prescribed treatment, and around 30% show no improvement even after several treatment attempts [12]. This highlights the need for more cost-effective, accessible and alternative treatments for depressive disorders. One of such intervention that has been suggested is physical exercise (E)/physical activity (PA) [13, 14]. It should be critically noted that many reviews do not define what is meant by E or PA and the terms are often used interchangeably [15–17]. It can be stated that exercise is always physical activity but physical activity is not necessarily exercise [18]. The key difference between the two terms is that exercise is planned and structured [15]. The American College of Sports Medicine (ACSM) defines PA as any physical activity that is produced by the contraction of skeletal muscle and results in a significant increase in caloric expenditure compared to energy expenditure at rest. PA was also included in the search in this review, as many studies use the terms synonymously [18].

It has been known for a long time that E and PA can reduce depressive symptoms and improve the quality of life of people with depression [1,13, 18]. E may also impact indicators of well-being in this population and may influence depressive symptoms through a variety of psychosocial and biological mechanisms [1, 4, 5]. Clinical practice guidelines in the USA, the UK, and Australia recommend PA as part of the treatment of depression. Despite the research and findings of recent years, it is still unclear what mechanism lies behind this and what type of E/PA is particularly beneficial [1, 5, 13, 19]. In 2014, Wegner et al. [1] published a paper to summarize the existing meta-analyses on the anxiolytic and depression-reducing effects of E [1]. The meta-analyses considered in Wegner’s paper consistently show a positive effect of E on anxiety and depression. The overall effect size found for the effect of E on depression was moderate, ES = 0.56 [1]. It was also shown that E is no less effective than pharmacological or psychotherapeutic treatments [1]. Regular physical activity (PA) can boost the levels of several growth factors in the brain, which supports neuronal plasticity and improves overall brain health. These benefits may help mitigate atrophic changes in the hippocampus. This indicates that PA could play a crucial role in both the prevention and treatment of brain-related disorders. [1].

More than 10 years have passed since the publication of these meta-analyses. Although these results provide evidence for the alleviation of depressive symptoms by E, these studies can be criticized for including studies of low methodological quality, such as quasi-experimental and cross-sectional studies [11, 13, 19, 20]. Careful consideration of the characteristics of meta-analytic studies revealed methodological aspects that could confirm previous assumptions. For example, the diagnosis of depression in a number of studies was not based on valid diagnostic criteria [8].

In addition, the way in which the subjects were recruited and whether they were clinical or non-clinical subjects seems to be decisive [21]. The problem here is that it is often not possible to prove the specific effects of the intervention used [22]. For example, the positive effects of meditation can also result from spending a lot of time with a carer, so the improvements in the meditation group could also be partly attributable to these non-specific effects (i.e. not directly to the meditation itself). It is therefore advisable to include a control condition for non-specific effects in non-inferiority studies that are matched in terms of time and attention [22]. It was found that such activities can also improve symptoms of depression. This raises questions about the actual antidepressant effects of this sham interventions and may lead to wrong conclusions [8, 20, 22].

Over the past 10 years, an increasing number of studies suggest that the relationship between PA and improvements in depressive symptoms may be influenced by mediators, and it is still unclear how much and what type of PA and E is beneficial [1, 5, 11, 13, 23]. The age of the patients could also influence the relationship between PA and depression [1, 18, 24]. Older depressed adults appear to show greater relief from depression through E [8].

Research of the last ten years showed that the positive effects of PA on depression cannot be explained by a single, isolated mechanism [1, 13]. Rather, the effects are most likely due to the interplay of several mechanisms that manifest themselves on a psychological (e.g. mood, sense of coping, self-efficacy) and neurobiological level (e.g. neurogenesis in the hippocampus, regulation of the hypothalamic-pituitary-adrenal axis, modulation of neurotransmitters) [1, 25–27]. Current research raises the question of whether the findings reported in the 2014 review by Wegner et al. [1] can be confirmed or whether more recent research provide new insights. The present work is intended as an update to the report by Wegner et al. [1].

### 1.1 Characteristics of depression

Depression is a serious mental disorder characterized by a high burden of symptoms [28]. This disorder can have serious consequences, including the possibility of suicidal thoughts or actions [10, 28]. Second-generation antidepressants are still the first choice in the treatment of depression [14]. Previous research has shown that many adults with depression go untreated and do not seek help for their symptoms [13]. Particularly in low- and middle-income countries, more than 75% of people do not receive appropriate support despite the existence of known and effective treatment methods for mental disorders [10]. This is still due to insufficient investment in mental health care, a lack of trained healthcare professionals and the social stigma associated with mental disorders [10]. During a depressive episode, a person experiences a depressed mood that manifests itself through sadness, irritability, or emptiness. In addition, they may lose pleasure or interest in activities [1, 29]. This episode is very different from normal mood swings as it lasts most of the day, almost every day, for a period of at least two weeks [10, 11]. Despite appropriate treatment, many patients still do not experience sufficient relief of their symptoms [30, 31]. Even though awareness of depression and its treatment options has increased since 2014, the problems described with conventional therapies such as antidepressants and psychotherapy show that there are still major challenges in terms of treatment options and effectiveness. A more accessible and accepted treatment method with fewer side effects would be desirable.

### 1.2 Prevalence Depression

Around 280 million people worldwide are affected by depressive disorders, making them one of the main causes of the global health burden [19]. Between 2015 and 2019, the prevalence of depression increased significantly, but without a corresponding increase in treatment [9]. In 2020, almost ten per cent of the American population and almost a fifth of adolescents and young adults were affected by depression within the last twelve months [9, 32]. An estimated 3.8% of the population is affected by depression, with 5% of adults (4% of men and 6% of women) and 5.7% of adults over the age of 60 belonging to this group [10]. Major depressive disorder (MDD) is the most common form of depression, with a lifetime prevalence of 6-15% [33]. This indicates an increasing trend since the work of Wegner et al. [1]. The average age of onset of major depressive disorder is 40 years, with 50% of all patients falling ill between the ages of 20 and 50 [5, 34]. Every year, over 700,000 people lose their lives to suicide, with suicide being the fourth most common cause of death for people aged 15 to 29 [10,11].

## 2 Methods

The studies by Wegner et al. [1, 18] were used as a guide for the methodology, as this current work is an update of the 2014 study conducted 10 years ago. However, this paper focuses exclusively on the effects of E/PA on depression. Unlike the paper from 2014 the effects of anxiety are not addressed here. All meta-analytic reviews for the present article were identified via databases such as PsycINFO, PsychARTICLES, PubMed, CINAHL Complete, SocINDEX, SPORTDiscus and Psyndex. Meta-analyses published after July 2014 were included only. The search terms used for the online resources were, “meta-analysis*, “depression” and “depressive disorder”, and “exercise” and “physical activity”. Only sources published in peer-reviewed journals in English were considered. The preliminary search yielded 1586 results with the keywords (meta-analysis or meta-analytic) AND (depression or depressive disorder) AND (exercise or physical activity or physical training). To obtain only relevant hits, the Boolean operator “title” was used for the search. The search process was as follows: TI (meta-analysis, meta-analysis) AND TI (depression or depressive disorder) AND TI (exercise or physical activity physical training). The search resulted in 288 hits. The literature search was completed on 15/05/2024.

### 2.1 Protocol and Registration

The protocol of this systematic review was registered on May 23, 2024 in PROSPERO (International prospective register of systematic reviews) at www.crd.york.ac.uk under the PROSPERO-ID CRD42024549007.

### 2.2 Eligibility Criteria

In order to assess the suitability of a meta-analysis for this article, the eligibility criteria listed in *Table 1* had to be met by the individual studies included. The criteria were structured according to the PICOS approach, which takes into account population, intervention, comparison group, results and study design [18, 35]. Only meta-analyses were considered, including longitudinal studies with control groups. The results of these studies are typically compared with a baseline value collected at the beginning of the study. This makes it possible to comprehensively understand the extent and direction of changes over time. [18].

**Table 1:** Eligibility criteria by category (PICOS).

| Category | Eligibility criteria |
| --- | --- |
| Population | Otherwise healthy adults with<br>a) a clinical diagnosis of depression diagnosed using clinical recognized diagnostic criteria or<br>b) depression or depressive symptoms assessed using any recognized diagnostic criteria |
| Intervention | Intervention consisting of physical exercise or physical activity |
| Comparator | Control group |
| Outcome | Benefits of physical exercise/activity<br>Effect size reported |
| Study design | Only meta-analyses were considered, including randomized controlled trials, cluster randomized controlled trials, controlled trials, longitudinal studies |

In contrast to Wegner et al. [1], participants with concomitant diseases such as cancer (or cancer survivors), chronic diseases, heart problems or postpartum depression were excluded from this study. Depression should be the main focus. The hormonal changes during pregnancy and childbirth can trigger strong changes in affective state [36, 37]. Due to the abrupt and dramatic changes in hormone levels after childbirth, some studies emphasise the role of hormonal factors in postpartum depression [38]. However, the extent to which these affective changes can be attributed to the altered hormone levels is still largely unknown [36]. These forms of depression were therefore excluded from this study.

In contrast to Wegner et al. [1], studies focussing only on children or older adults were excluded. Since an Umbrella Review on the effects of exercise on depression in older adults was published in 2016 and a “Systematic Review of Meta-Analyses on Exercise Effects on Depression in Children and Adolescents” in 2020, only studies in which at least 80% of the participants were between 18 and 59 years old were included in this study [18, 39].

### 2.3 Data Analysis

In all included meta-analyses, the standardized mean difference (SMD = M1 - M2/SDpooled) was used as a measure of effect size. The reported effect sizes of each included meta-analysis were pooled to calculate and discuss an overall effect size [1]. If the calculation of the effect size in a study deviates from this measure, the respective estimate may have to be converted. However, Hedges’ g and the standardised mean difference (SMD) are equivalent in many contexts, especially if the sample size is large enough. Therefore, the one study [8] that specified an effect size corrected according to Hedges’ g did not need to be converted [40, 41].

### 2.4 Quality Assessment

To assess the methodological quality of the included systematic reviews with meta-analyses, two researchers independently completed the AMSTAR-2 checklist (A Measurement Tool to Assess Systematic Reviews) [18, 42].

The final agreement between the two independent researchers on the results obtained with the AMSTAR 2 checklist regarding the methodological quality of the meta-analyses is as follows: 14, 30, 43 = *high quality review*; 8, 44 = *moderate quality review*; 5, 27 = *low quality review*; 23, 45-47 = *critically low quality review*.

## 3 Results

### 3.1 Study Selection

The literature search identified a total of 1,586 studies that addressed systematic reviews with meta-analyses in this area. After duplicates were removed, two independent researchers screened 1,298 titles and abstracts. After a joint evaluation, 68 studies remained that were classified as potentially relevant and examined in full text. Eleven studies met the eligibility criteria and were included in this review (*for more information see the Flow chart of the selection process, Figure 1*).

**Figure 1:**
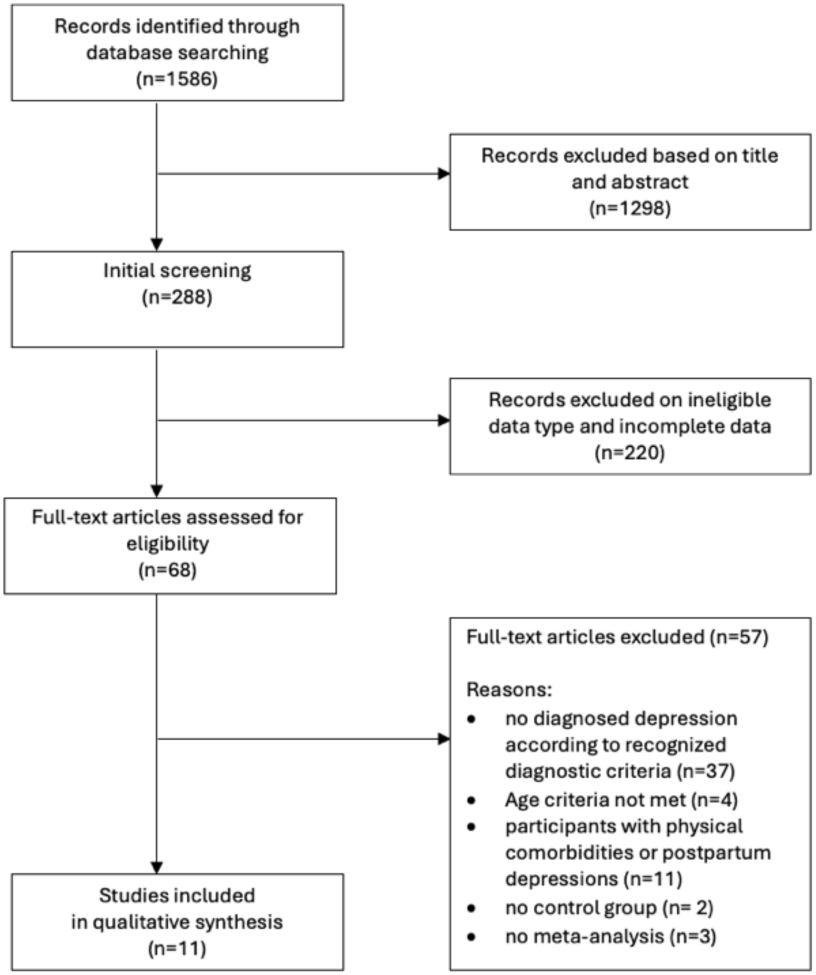
Flow chart of the selection process.

### 3.2 Characteristics of the Studies

Meta-analyses were included if they examined either randomized controlled trials (RCT), cluster randomized controlled trials, controlled trials, or longitudinal studies. Eleven meta-analytic reviews with reported effect sizes were found on the effect of exercise and physical activity on depression. A total of 16.255 participants and 229 individual studies were included in all reviewed analyses. All articles focused exclusively on randomised controlled trials. Of the eleven articles, all dealt with patients with depression and two reviews included only women. The general characteristics information are summarized *in Table 2*.

**Table 2:**
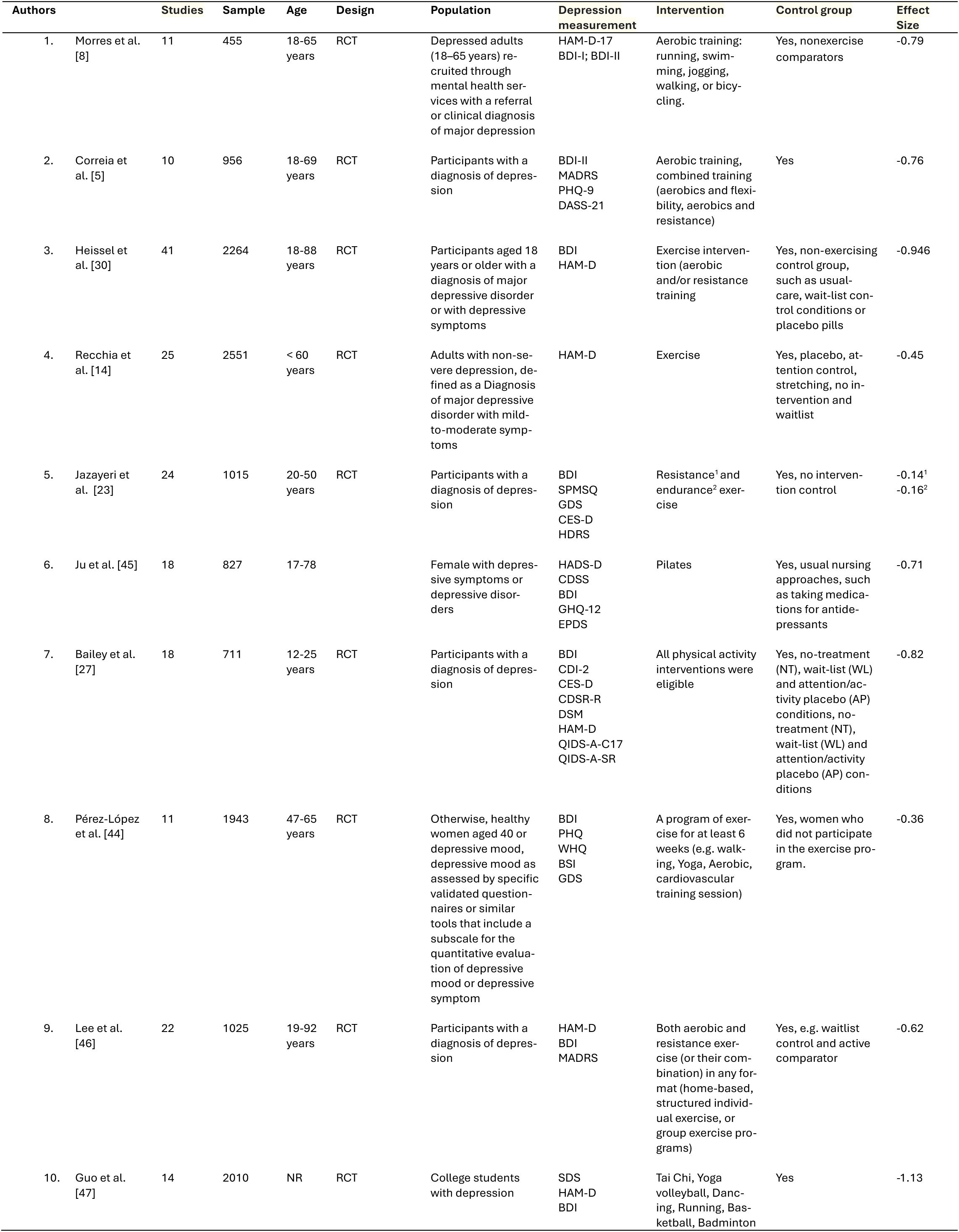
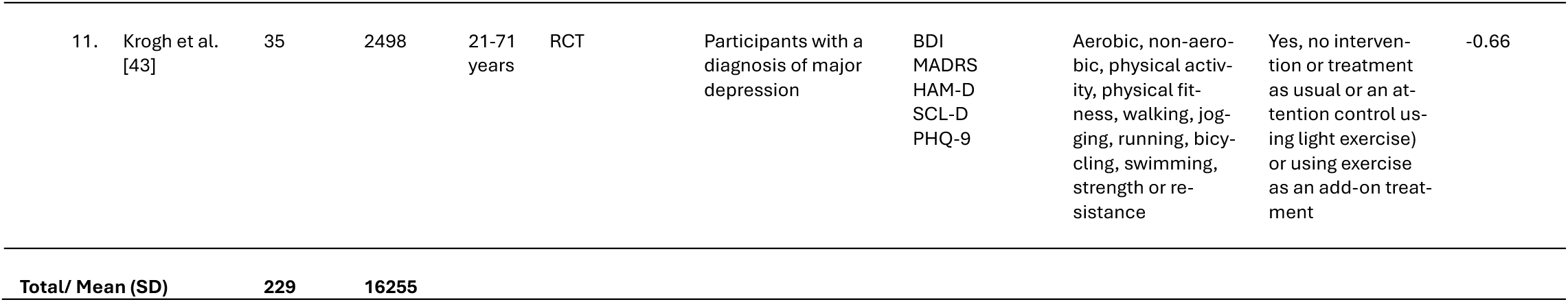
General characteristics of included meta-analyses.

| Authors | Studies | Sample | Age | Design | Population | Depression measurement | Intervention | Control group | Effect Size |
| --- | --- | --- | --- | --- | --- | --- | --- | --- | --- |
| 1. Morres et al. [8] | 11 | 455 | 18-65 years | RCT | Depressed adults (18–65 years) recruited through mental health services with a referral or clinical diagnosis of major depression | HAM-D-17<br>BDI-I; BDI-II | Aerobic training: running, swimming, jogging, walking, or bicycling. | Yes, nonexercise comparators | -0.79 |
| 2. Correia et al. [5] | 10 | 956 | 18-69 years | RCT | Participants with a diagnosis of depression | BDI-II<br>MADRS<br>PHQ-9<br>DASS-21 | Aerobic training, combined training (aerobics and flexibility, aerobics and resistance) | Yes | -0.76 |
| 3. Heissel et al. [30] | 41 | 2264 | 18-88 years | RCT | Participants aged 18 years or older with a diagnosis of major depressive disorder or with depressive symptoms | BDI<br>HAM-D | Exercise intervention (aerobic and/or resistance training) | Yes, non-exercising control group, such as usual-care, wait-list control conditions or placebo pills | -0.946 |
| 4. Recchia et al. [14] | 25 | 2551 | < 60 years | RCT | Adults with non-severe depression, defined as a Diagnosis of major depressive disorder with mild-to-moderate symptoms | HAM-D | Exercise | Yes, placebo, attention control, stretching, no intervention and waitlist | -0.45 |
| 5. Jazayeri et al. [23] | 24 | 1015 | 20-50 years | RCT | Participants with a diagnosis of depression | BDI<br>SPMSQ<br>GDS<br>CES-D<br>HDRS | Resistance<sup>1</sup> and endurance<sup>2</sup> exercise | Yes, no intervention control | -0.14<sup>1</sup><br>-0.16<sup>2</sup> |
| 6. Ju et al. [45] | 18 | 827 | 17-78 |  | Female with depressive symptoms or depressive disorders | HADS-D<br>CDSS<br>BDI<br>GHQ-12<br>EPDS | Pilates | Yes, usual nursing approaches, such as taking medications for antidepressants | -0.71 |
| 7. Bailey et al. [27] | 18 | 711 | 12-25 years | RCT | Participants with a diagnosis of depression | BDI<br>CDI-2<br>CES-D<br>CDSR-R<br>DSM<br>HAM-D<br>QIDS-A-C17<br>QIDS-A-SR | All physical activity interventions were eligible | Yes, no-treatment (NT), wait-list (WL) and attention/activity placebo (AP) conditions, no-treatment (NT), wait-list (WL) and attention/activity placebo (AP) conditions | -0.82 |
| 8. Pérez-López et al. [44] | 11 | 1943 | 47-65 years | RCT | Otherwise, healthy women aged 40 or depressive mood, depressive mood as assessed by specific validated questionnaires or similar tools that include a subscale for the quantitative evaluation of depressive mood or depressive symptom | BDI<br>PHQ<br>WHQ<br>BSI<br>GDS | A program of exercise for at least 6 weeks (e.g. walking, Yoga, Aerobic, cardiovascular training session) | Yes, women who did not participate in the exercise program. | -0.36 |
| 9. Lee et al. [46] | 22 | 1025 | 19-92 years | RCT | Participants with a diagnosis of depression | HAM-D<br>BDI<br>MADRS | Both aerobic and resistance exercise (or their combination) in any format (home-based, structured individual exercise, or group exercise programs) | Yes, e.g. waitlist control and active comparator | -0.62 |
| 10. Guo et al. [47] | 14 | 2010 | NR | RCT | College students with depression | SDS<br>HAM-D<br>BDI | Tai Chi, Yoga volleyball, Dancing, Running, Basketball, Badminton | Yes | -1.13 |

|  |  |  |  |  |  |  |  |  |  |  |
| --- | --- | --- | --- | --- | --- | --- | --- | --- | --- | --- |
| 11. | Krogh et al. [43] | 35 | 2498 | 21-71 years | RCT | Participants with a diagnosis of major depression | BDI<br>MADRS<br>HAM-D<br>SCL-D<br>PHQ-9 | Aerobic, non-aerobic, physical activity, physical fitness, walking, jogging, running, bicycling, swimming, strength or resistance | Yes, no intervention or treatment as usual or an attention control using light exercise) or using exercise as an add-on treatment | -0.66 |
| Total/ Mean (SD) |  | 229 | 16255 |  |  |  |  |  |  |  |

### 3.3 Meta-Analytical Findings for the Effect of E/PA on Depression

Recently, several authors have conducted comprehensive meta-analyses to investigate the effects of E/PA on depression, examining various factors that may moderate this effect [1, 18]. A total of eleven meta-analyses on the depression-reducing effect of E and PA were included in this review. The results of these analyses are summarized below. The effect of intervention versus control was a SMD of -0.61 (95% CI [− 0.78; -0.43], *p* = <0.01). Effect size calculations can be found in the forest plot *(Figure 2)*.

**Figure 2:**
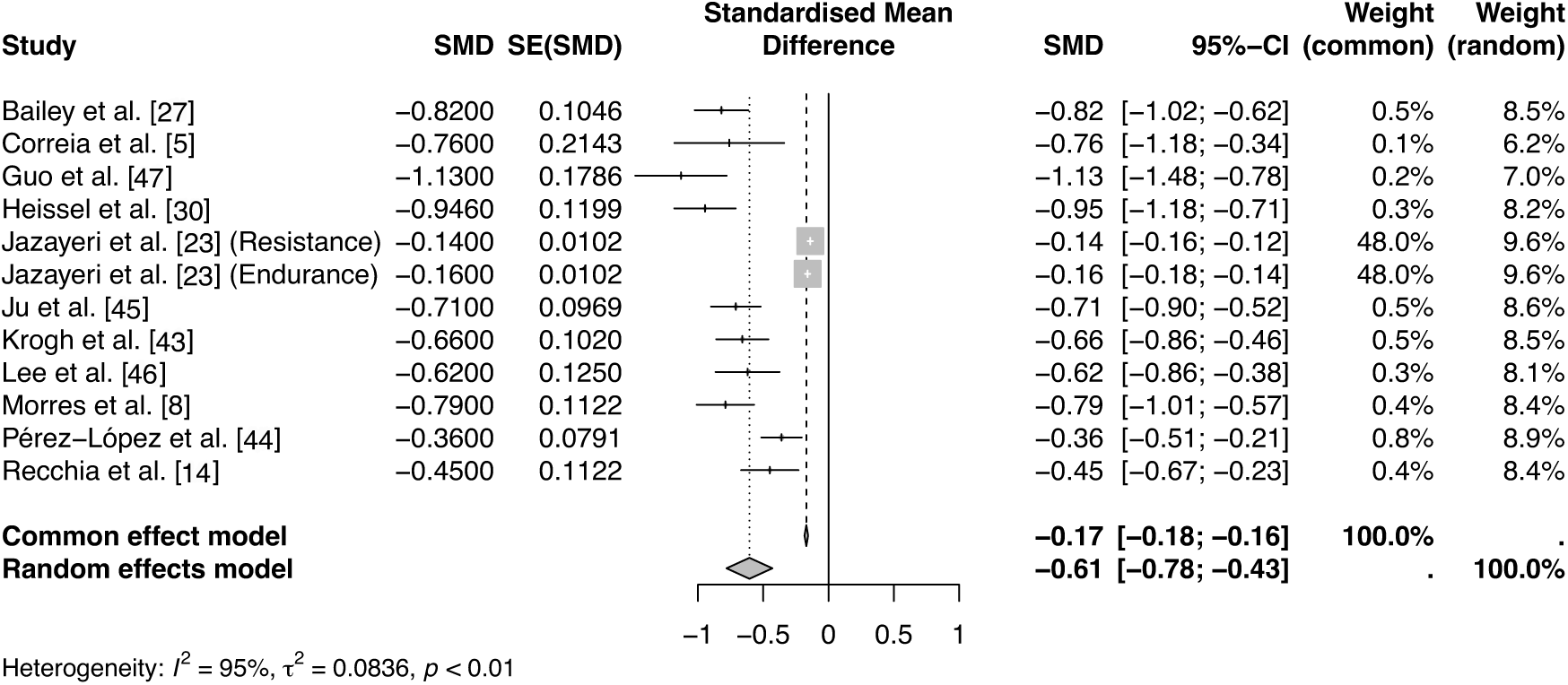
Forest plot.

#### Randomized controlled trials (RCT)

In contrast to Wegner et al. [1], only meta-analyses with randomized controlled trials are included in the present study. Wegner et al. [1], reported that the meta-analyses that tended to use randomized controlled trials had higher effect sizes (ES = -0.65) than studies that did not exclusively use randomized controlled trials (ES = -0.45). This must be taken into account when discussing the results of the present study. The ES determined here (-0.61) is therefore more comparable with the ES (-0.65) of the studies in Wegner et al. [1], comparing the randomized-controlled-trials effect to less rigid study designs.

#### Measures of depression

In numerous studies, depression is diagnosed according to DSM or ICD-10 criteria [43]. The Beck Depression Inventory (BDI) and the Hamilton Depression Scale (HAM-D) are the most frequently used questionnaires [30]. Other commonly used instruments to measure depression include the Center for Epidemiologic Studies Depression Scale (CES-D) and the Hamilton Depression Rating Scale (HAM-D) [27]. *For a detailed list, see Table 2*.

#### Comparison with other therapies

Research to date shows great variability in the minimum duration, intensity, and type of PA/E in its effect on depressive symptoms [1]. While some studies investigated the general effect of PA on depression other analyses focused on the direct effect of specific chronic E on depression.

Physical activity is defined here as planned, structured, repetitive and purposeful physical activity to improve or maintain physical fitness [14, 18, 30]. There was a moderate to large significant effect of E compared to no intervention (ES = -0.76 to -1.24) [5, 8, 30]. In addition, E had a significant effect compared to usual care (ES = -0.48 to -0.82) (usual care, waiting list control conditions, or placebo pills) [27]. Back in the days, there was no consensus as to whether physical exercise has a greater impact on the symptoms and cognitive functions of depression compared to taking antidepressants [48]. Recchia et al. [14] observe the efficacy of E, antidepressants, and combined treatments on depressive symptoms in adults with non-severe depression. The results suggest that all treatments had similar positive effects on depressive symptoms compared to the control groups, with no treatment being superior to the other [14]. Wegner et al. [1] also found no benefit of exercise compared to psychosocial interventions, low-intensity exercise, or relaxation interventions. Lee et al. [46], on the other hand, report that exercise as an adjunct to standard treatment (e.g. pharmacotherapy, psychotherapy, electroconvulsive therapy; ECT) has a moderate antidepressant effect compared to standard treatment alone. They also provide new evidence that exercise in combination with standard treatment has a significant moderate antidepres-sant effect compared to standard treatment alone. Furthermore, Lee et al. [46] report that the addition of exercise to standard treatment did not have a negative effect on acceptability. Some studies have even shown that the combination of exercise and standard treatment improves medication adherence and compli-ance with treatment protocols. In addition, the results suggest that exercise positively influences the response to treatment and the course of symptom worsening [46]. Wegner et al. [1] reported in their paper that several authors were unable to find any difference between exercise programs and cognitive therapy in de-pressed patients. These results can be confirmed in the present study [8].

In summary, what Wegner et al. [1] reported can be confirmed in the present updated analysis. The effect of E/PA on depression is not demonstrably beneficial compared to other interventions. However, it has been shown to be no less effective, even when compared to antidepressant medication or psychotherapy. There seems to be a consensus that E/PA is superior to no treatment and the combination of E with conventional therapies seems to have a superior effect.

#### Type of exercise

Morres et al. [8] focused exclusively on the effect of aerobic E (e.g. running, swimming, jogging, walking, or bicycling) on depression in their study. In this review, E was found to have a significantly large antidepressant effect in adult patients recruited through mental health services who had a referral or clinical diagnosis of major depression. Compared to antidepressants, it showed a strong antidepressant effect [8]. Similar effects were observed when the effect of aerbobic E was compared with psychological treatments, either as monotherapy or as part of a multi-therapeutic program. What is also worth mentioning is that aerobic E was preferred over psychological treatments or antidepressants/TAU [8]. There are also other reviews that report the superior effect of aerobic E compared to other forms of E such as resistance or flexibility training [5, 30].

Some reviews excluded studies that used yoga, tai chi or other mental/physical activities [14, 30]. The reason for this is that these mind-body interventions focus on behavioral techniques. These include deep breathing, meditation/mindfulness, and self-awareness, among others, and thus a range of behavioral techniques that could interfere with the effect of physical E [14, 30] or should be studied with a sham condition [22]. Guo et al. [47], on the other hand, compared seven different exercise interventions, including tai chi. The results showed that tai chi and yoga reduced depression in students more than badminton or basketball [47]. Jazayeri et al. [23] focused on the effect of endurance and resistance training on depression. On the one hand, they found that both types of training were effective in reducing depression compared to the control group; on the other hand, there was no difference in the effectiveness of the two interventions. Both reduced depressive symptoms equally [23]. Heissel et al. [30] and Correia et al. [5] agree that chronic aerobic exercise as a single intervention has a greater effect than mixed interventions (e.g. in combination with resistance training). Recchia et al. [14] also report that a combination therapy has no greater positive effect on depressive symptoms than either treatment alone. This is in contrast to the findings of Wegner et al. [1], who report a superior effect of combination training.

In summary, several studies concluded that moderate-intensity interventions have a more positive effect compared to high-intensity interventions [5, 14, 30, 44]. Only Krogh et al. [43] report on the superior effect of high-intensity/high-dose exercise. In addition, group interventions appear to be preferable to individual training sessions [5, 30, 46]. It can be concluded that participants seem to benefit more from interventions with a shorter weekly duration [30, 43].

#### Gender

In general, women and men appear to benefit from E to reduce their depressive symptoms, but women are more likely to suffer from depression [5, 10, 44]. The reasons for this are not clear [44]. In a meta-analysis that focused exclusively on women, it was found that programmed exercise significantly reduced depressive symptoms in middle-aged to older women (47 to 65 years). The benefit appeared to be independent of whether the training intensity was low or moderate [44]. Ju et al. [45] found a significant improvement in depressive symptoms in women as a result of Pilates. However, this was not a comparative study; the effect of Pilates on depressive symptoms in men was not investigated [45].

### 3.4 Neurobiological Explanations

Taking a closer look at the neurobiology of depressed people, dysfunction of the hypothalamic-pituitary-adrenal (HPA) axis, increased secretion of corticotropin-releasing hormone (CRH), decreased responsiveness to glucocorticoids, and increased pituitary size and activity, among others, can be observed [1, 49]. Despite 10 years have past it is still (and will properly remain impossible) to account the positive effect od E and PA to one neurologoical change. The exact mechanisms underlying the antidepressant effects of PA in MDD are still unclear, speculative, and largely based on animal studies or results from studies conducted in humans without MDD [31, 50–52]. On this basis, the efficacy of PA and E in depressive disorders is attributed to physiological changes in monoamine metabolism, hypothalamic-pituitary-adrenal (HPA) axis function, neurotrophic factors and neuroinflammation, among others [1, 49, 50].

It is important to note a still important cause of depression thought to be a dysfunction of the serotonin system (5-hydroxytryptamine; 5HT) [1, 12]. Conventional anti-depressants for the treatment of depression are typically designed to block the reuptake or degradation of monoamines such as 5-hydroxytryptamine (5-HT or serotonin) and noradrenalin (NA). Selective serotonin reuptake inhibitors (SSRIs) are still the most commonly prescribed drugs for the treatment of depression [12, 50]. Nevertheless, depressive disorder is a complex disease with changes in several biomarkers that go beyond monoamines. The cause of depression is not a simple deficiency of key monoamines alone [50]. Therefore, it is not surprising that treatment with common antidepressants, which mainly target monoamines, often shows limited therapeutic effect [50]. Various systematic reviews have described the influence of PA on the serotonergic system and the function of the HPA axis [53]. It is assumed that PA could improve the efficiency of the response of the monoamine system and normalize the HPA axis in depressed patients [1, 50].

#### HPA axis activity and cortisol

Natural stress, whether physical or psychological, activates two main response systems in the body: the sympatho-adrenomedullary system, which triggers an alarm and increases the heart and respiratory rate, and the hypothalamic-pituitary-adrenal axis (HPA axis), which releases cortisol, a key hormone for stress reduction [1, 54]. Depressed patients often show changes in the HPA system, leading to impaired regulation and an increase in cortisol in the blood [1, 53, 55]. These changes can contribute to hyperactivity of the HPA axis, which in turn can exacerbate the symptoms of depression. Overall, research suggests that HPA axis activity plays an important role in the regulation of mood and that dysregulation of this system may contribute to the development of mental disorders [1, 53]. This suggests that the relationship between the HPA axis and cortisol levels could play an important role in the development and progression of depression.

The HPA axis responds to environmental stress by activating the release of serotonin, noradrenaline, and dopamine from various brain regions such as the amygdala and the hippocampus [53]. This leads to the synthesis of corticotropin-releasing hormone in the hypothalamus, which binds to receptors in the anterior lobe of the hypophysis and triggers the release of adrenocorticotropin (ACTH) into the blood-stream. ACTH in turn stimulates the adrenal glands to produce and release cortisol [12, 53]. To maintain homeostasis, there are negative feedback mechanisms in which the glucocorticoids bind to glucocorticoid receptors (GR) to inhibit the further release of corticotropin-releasing hormone. In people with major depressive disorder (MDD), the sensitivity of these glucocorticoid receptors is impaired, resulting in reduced negative feedback. The result is excessive secretion of the corticotropin-releasing hormone and increased production of glucocorticoids such as cortisol [12]. Acute physical exertion is a direct stressor that affects hormone production, including cortisol. Cortisol levels typically rise approximately 20 to 30 minutes after the termination of stress [56, 57]. However, chronic stress can lead to pathological changes if the organism is unable to end the stress response [54, 58]. This also applies to mental illnesses such as depression, whose pathophysiology is closely linked to the endocrine stress system. In addition, studies suggest that increased chronic PA promotes the lowering of catabolic cortisol [59] and the neuroplasticity of certain brain structures [1, 58]. This is associated with an improvement in synaptogenesis, neurogenesis, the release of neurotrophins, and neuroendocrinological changes. The changes may be associated with benefits for cognitive and affective functions and this may translate into a reduction in psychosocial stress symptoms in the long term [52]. However, these results should be interpreted with caution due to the small number and heterogeneity of the studies [59]. In addition, the study found a large reduction in cortisol levels in MDD individuals for training at a frequency of five times per week, while there was a moderate effect at three times per week and a small effect at twice per week [59]. Fernandes et al. [51] also investigated the decline in cortisol levels in depressed people. One group received only antidepressants, while the other group also completed aerobic training. Over a period of four weeks, it was observed that cortisol levels decreased regardless of whether or not sertraline was combined with E. It was also found that clinical remission was achieved more frequently in the group that combined antidepressants and PA [51]. Although the study did not detect changes in cortisol levels or interleukins in people with MDD when PA was used as an adjunct to antidepressants, this research suggests that cortisol is the biomarker most sensitive to antidepressant treatment [51]. The most important physiological factor determining the neuroendocrine stress response appears to be the intensity and duration of activity [60]. Therefore, E should be differentiated into acute E (one-time exercise) and chronic E (repeated exercise or exercise training). This means that chronic endurance training can be used both as a preventative and therapeutic instrument to reduce the level of cortisol [54]. A study comparing different chronic interventions indicates that coordinative training can lead to a greater reduction in cortisol concentration and stress levels compared to chronic endurance training [61]. However, other results suggest that not all people suffering from depression show a change in the HPA axis. Therefore, not everybody would benefit from treatment that specifically targets components of the HPA axis [12].

#### Monoamine levels

Studies still show that people suffering from depression often show reduced levels of 5-hydroxyindoleacetic acid (5HIAA), the most important metabolite of 5HT (serotonin). In addition, reduced tryptophan levels in plasma, a low tryptophan-to-amino acid ratio, and abnormalities in 5HT function are frequently observed [1, 53]. Carneiro et al. [50] investigated the influence of chronic PA as monoamine responses in women with depressive disorders. It was assumed that the training group showed a significant increase in monoamine levels compared to the control group. Surprisingly, the training group showed a slight decrease in dopamine levels compared to the control group. The decrease in serotonin levels in the training group contradicts the hypothesis that PA could cause an increase in serotonin levels and thus have a positive effect on depressive symptoms. These results suggest that the biological mechanisms mediating the effects of E on depressive disorders are more complex than previously thought [50].

#### Adaptation in neuronal structures

The brain-derived neurotrophic factor (BDNF) is a neurotrophin that plays a central role in the regulation of neuronal plasticity of synaptic activity, hippocampal function, and the learning process, among others [1, 33, 52]. BDNF expression can be stimulated by serotonin [53]. There is a link between low circulating levels of BDNF and affective/emotional dysregulation as seen in depressive disorders [33, 62]. BDNF is a potential biomarker for the success of depression therapy, as research increasingly suggests that BDNF may play a role in improving depressive behaviors through E [1, 31, 52]. Studies conducted with outpatients with MDD and persistent depression showed that both acute and regular E caused an increase in BDNF [33, 52]. Higher BDNF levels are associated with better cognitive and psychiatric status in both depressed patients and healthy individuals [53]. The improvement in the biomarker caused by E is associated with a decrease in depressive symptoms [33]. Previous studies have concluded that both endurance training and short-term high-intensity anaerobic exercise can lead to an increase in BDNF levels in both healthy and depressed patients [53, 63, 64].

Murawska-Ciałowicz et al. [65] investigated the effects of nine weeks of training with four different types of high-intensity on the resting concentration of BDNF. The results showed that the resting concentration of BDNF did not change significantly or was even reduced [65]. One meta-analysis found that chronic aerobic exercise in individuals with MDD was not associated with an increase in resting concentrations of BDNF in the periphery blood [66]. The meta-analysis by Kurebayashi and Otaki reports that E did not significantly increase BDNF levels in patients with MDD [67]. Although the study by Szuhany and Otto [68] confirms that physical activity leads to a significant increase in BDNF levels in both depressed and healthy adults, no significant correlation was found between changes in BDNF levels and improvement in depressive symptoms. One study investigated the effects of aerobic exercise on BDNF and cortisol levels in people with depression. However, it was not possible to establish an interaction between aerobic training and the concentration of BDNF and cortisol. The reasons for this are speculative [69]. Therefore, there is some evidence to suggest that BDNF may not be the best or only biomarker for the positive effect of exercise on depression [66, 68].

#### Plasticity

Wang et al. [62] report that six months of moderate-intensity aerobic training leads to a significant increase in the volume of grey matter in the prefrontal cortex in patients with depression. Reduced grey matter volume is an important physiological sign of depression. The increase was directly proportional to the amount of training. One study has shown that acute aerobic exercise can cause changes in brain connections in people with depression, particularly between the prefrontal cortex and the temporal cortical region associated with mood regulation [70]. The results showed that both moderate endurance training and high-intensity interval training improved the mood of the participants. This improvement in mood was associated with a stronger influence of the prefrontal cortex compared to the temporal cortical regions [70].

#### Cannabinoids

An alternative explanation gaining more and more interests is that during moderate PA, the concentrations of circulating endocannabinoids increase in people with depressive disorders. Endocannabinoids are lipids involved in processes such as pain, mood and memory and are associated with the acute affective responses during physical activity in people with depression [71, 72]. Numerous studies point to the role of cannabinoid receptors in influencing mental illnesses such as depression. There is several evidence that endocannabinoid levels are disturbed in depression [73]. Cannabinoid receptor activation leads to depression-like behavior [74]. On the other hand, CB1 cannabinoid receptor antagonists show antidepressant effects. Mice lacking the CB1 cannabinoid receptor exhibit abnormal behavioral patterns associated with mood disorders. These behavioral changes include impaired molecular functions and dysregulation of neurotransmitter systems associated with depression [73]. Experimental and clinical studies analyzing the endocannabinoids AEA (anandamide) and 2-AG (2-arachidonoylglycerol) have shown that these lipids are present at lower levels in people with depression. This is consistent with pharmacological studies indicating that modulation of the endocannabinoid system by enhancers or inhibitors of AEA or 2-AG may have positive therapeutic effects in mental disorders [73]. Thus, it can be hypothesized that pharmacological manipulation of endogenous endocannabinoid levels represents a new possibility for various medical treatments for depression. However, studies are needed to understand the exact mechanism of action by which the endocannabinoid system modulates depression [74].

## 4 Discussion

### 4.1 Summary of evidence

The aim of this article was to update the findings by Wegner et al. [1]. For this purpose, meta-analyses focusing on the effects of physical activity in adults on measures of depressive outcomes were reviewed. The overall effect size of the included meta-analyses was medium regarding the alleviation of depressive symptoms by E/PA. The effect of intervention versus control was SMD of = -0.61 (95% CI [-0.78; -0.43], *p* = <0.01).These results are comparable to those of Weger et al. [1]. They reported a moderate overall effect size for the effect of exercise on depression (ES = -0.56). As mentioned above, the ES was slightly higher for studies that focused exclusively on randomized controlled trials (ES = -0.65). Similar results were also observed in children (d = -0.50) [18]. In older adults, an umbrella review reported a small effect in favor of exercise on depression [39]. Given these results, E/PA can be considered a relevant treatment option for depression in all age groups.

Due to the limited and inconsistent E types, E duration and E frequency, the dose-response relationships are difficult to discern [5]. However there was a difference between 2014 and now. Wegner et al. [1] reported that either combined exercise programs or strength, weight, or resistance training alone were more effective than aerobic exercise. The most common intervention used in this work was aerobic exercise. Positive effects were consistently found for this intervention type. However, other types of exercise such as endurance and strength training also provide promising results [23, 30]. Wegner et al. [1] came to the conclusion that the results regarding the dosage of the exercises varied greatly and do not allow any conclusive recommendations to be made for practice. The optimal duration and frequency of physical interventions is still not clear. However, the average duration for which a positive effect of exercise has been observed appears to be around 45 minutes, three times a week, at moderate intensity [5, 8, 14, 27, 45]. Various meta-analyses and systematic reviews have shown that physical exercise can be similarly effective as psychotherapy and pharmacotherapy in the treatment of mild to moderate depression [5, 30]. This has also been reported by Weger et al. [1, 18]. The antidepressant effect of PA/E could be used as an inexpensive and easy-to-implement measure for the prevention and treatment of depression [19]. Furthermore, physical exercise is a less stigmatizing treatment option for people with depression. The importance of rapid access to treatment is underlined by the fact that prognosis worsens as the duration of depression increases. In addition, more than half of people with depression worldwide receive no treatment at all [31].

In order to implement physical interventions effectively, a well thought-out exercise and training concept is crucial. Exercise and training must be individually adapted and controlled, similar to medication. Due to the different physiological responses to physical interventions, which depend on many factors, individualization is important. This approach aims to maximize the effectiveness of the intervention by taking into account the differences between individuals [75]. Physical interventions should be prescribed and monitored taking into account specific markers of internal stress. Adjustment of internal loading requires careful and individualized adjustment of external loading. The optimal exercise and training prescription is likely to emerge from empirical evidence and interdisciplinary collaboration that integrates the perspectives of patients, medical experts, sports scientists, and practitioners [76].

Ten years have passed since the publication of Wegner et al. [1] and it is still not possible (and will remain so) to attribute the positive effect of E and PA to specific neurobiological changes. The precise mechanisms by which physical activity exerts its antidepressant effects in major depressive disorder remain uncertain, speculative, and primarily grounded in research involving animal models or studies conducted with individuals not suffering from MDD [1, 31, 50–52].

In summary, research shows that physical exercise has a consistently positive, moderate effect on depressive symptoms in adults without causing significant side effects. This confirms previous findings that PA/E should be considered as a valuable complementary therapy for the treatment of mental health problems in people of all ages [1, 18]. However, further research in this area is needed. Future studies should further explore the effects of these exercise interventions to support the development of targeted medical interventions and to determine the optimal dose-response relationship.

### 4.2 Strengths and limitations

It should be noted that only two of the eleven meta-analyses were of high quality. While six of the reviews were rated as low or even critically low *(see section 2.6 Quality assessment*). It should also be noted that the effects found in favor of exercise shrink when the analyses are limited to studies with a low risk of bias [30]. Krogh et al. [43] reported that studies with less than high risk of bias yielded significantly lower effect estimates for the impact of exercise on depression. This suggests that exercise interventions have only small antidepressant effects, depending on how much of the effect is due to bias and how much is due to the intervention. However, Recchia et al. [14] restricted their analyses to studies with a low risk of bias and were still able to demonstrate a medium effect (SMD = -0.66). Consequently, there is a need for more robust, rigorous and high quality studies to allow a realistic comparison of data between meta-analyses. On a positive note, all included meta-analyses were based on randomized controlled trials. The importance of conducting randomized controlled trials to investigate the effects of interest lies in their high quality [18, 77, 78].

## Data Availability

All data produced in the present work are contained in the manuscript

## 6 Appendix

**Supplemental 1:**
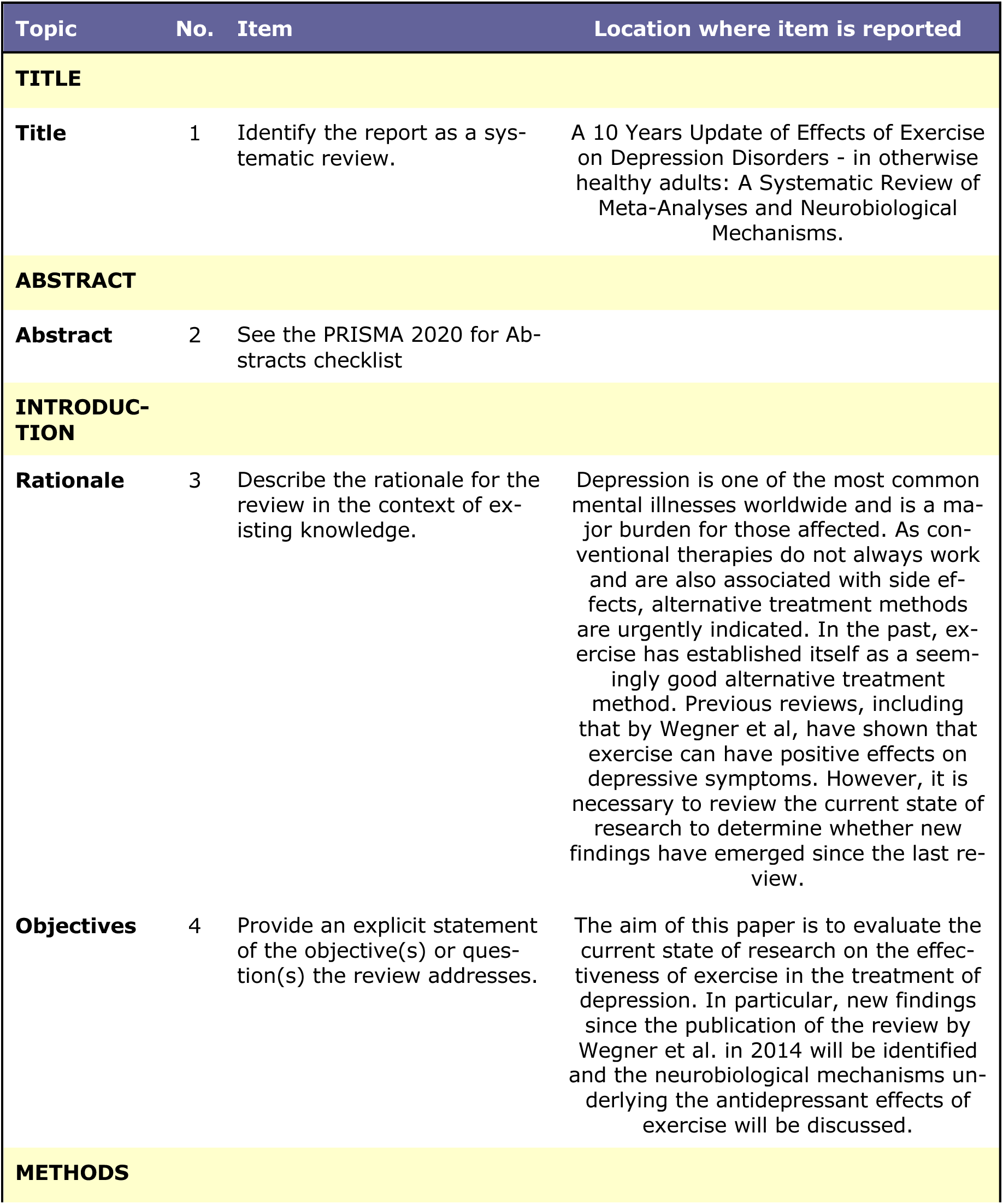
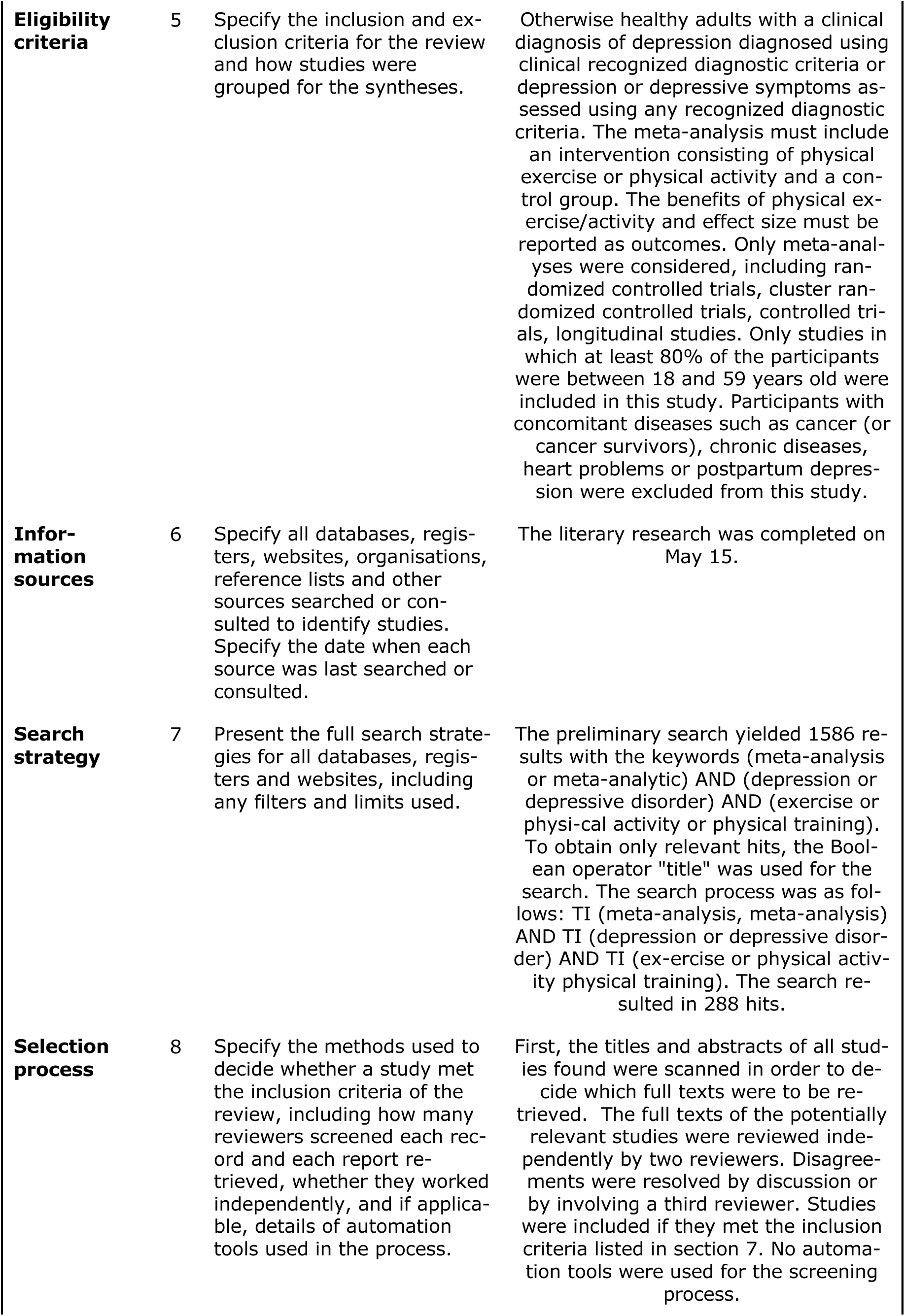
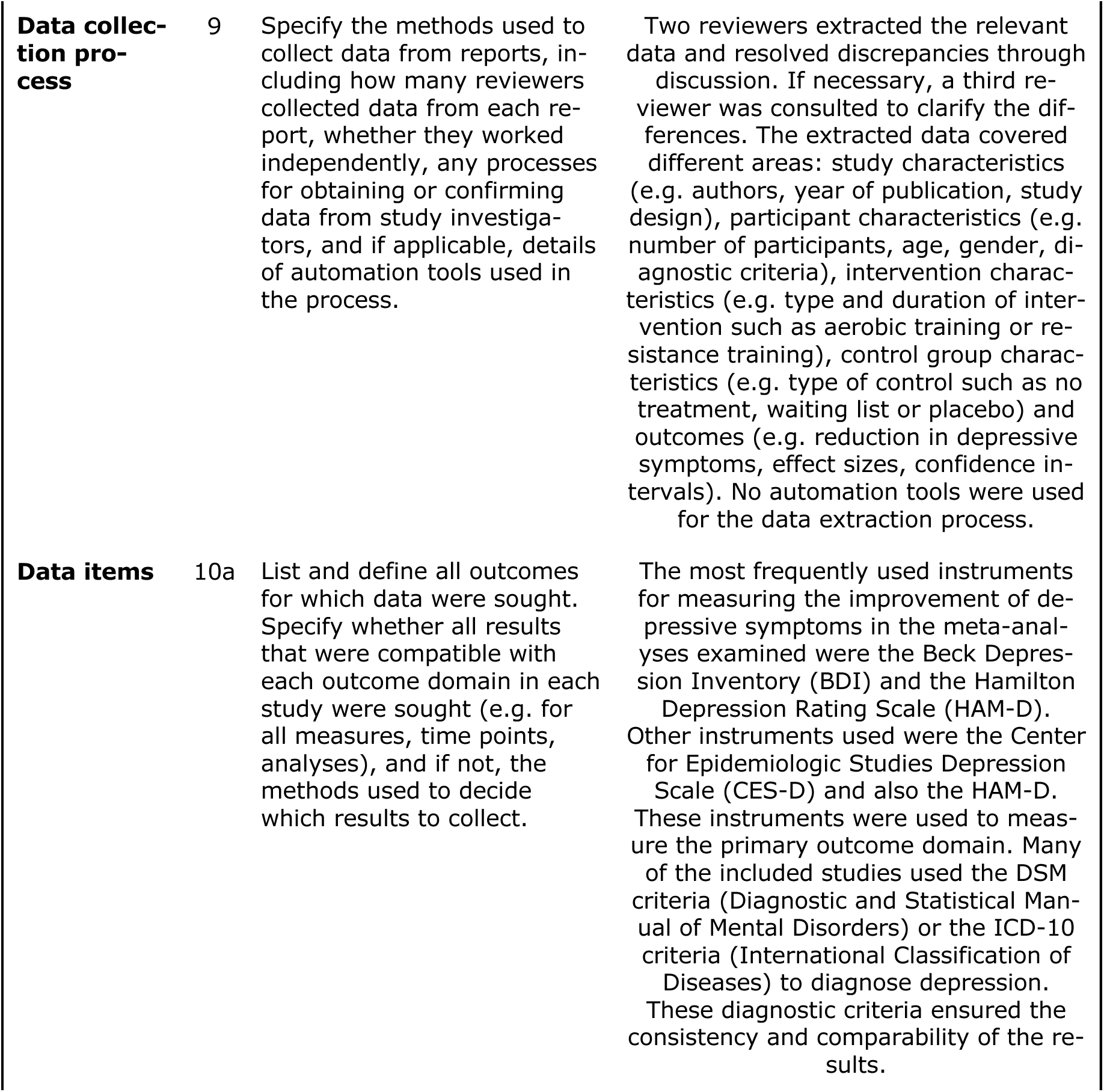
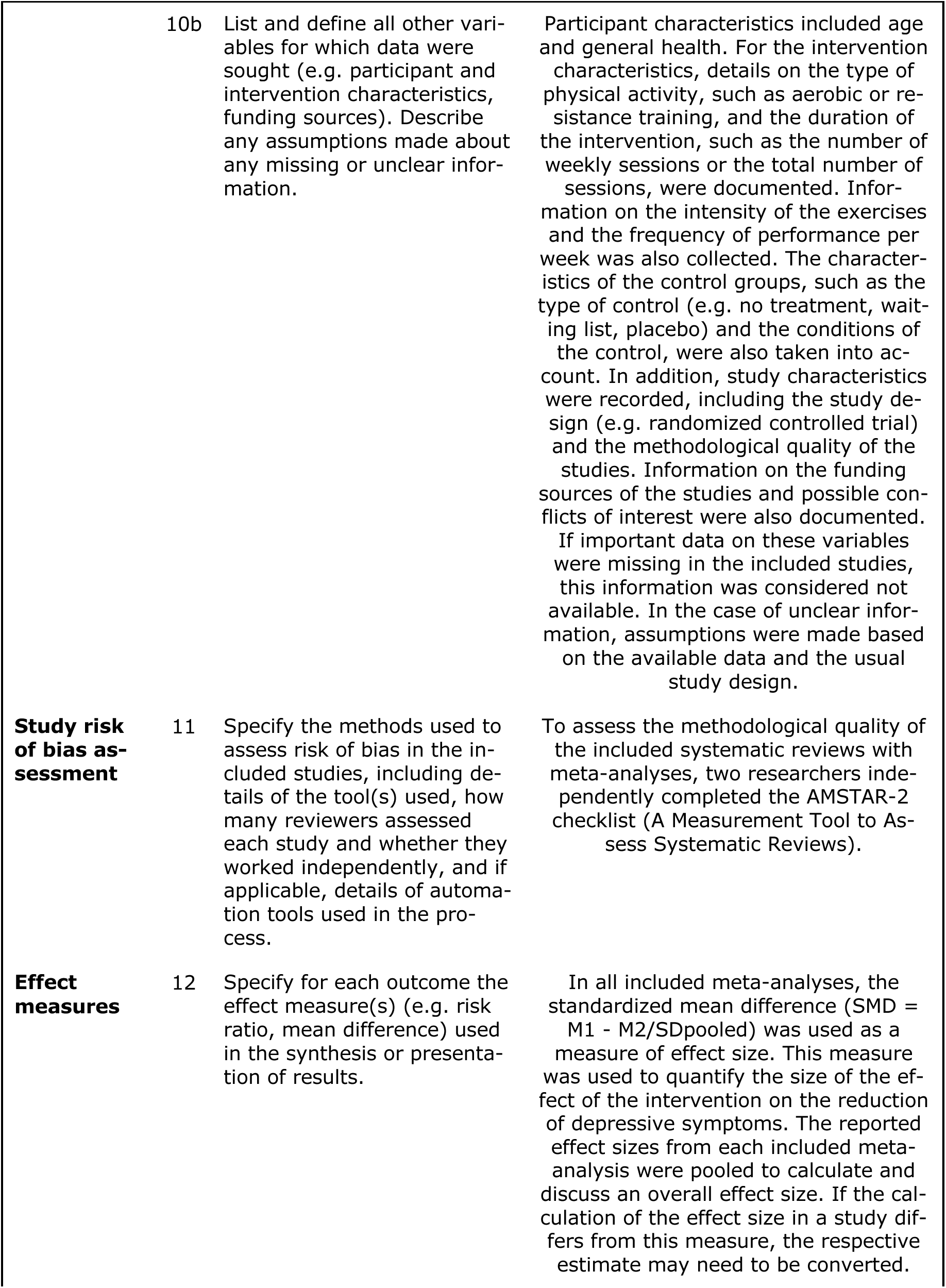
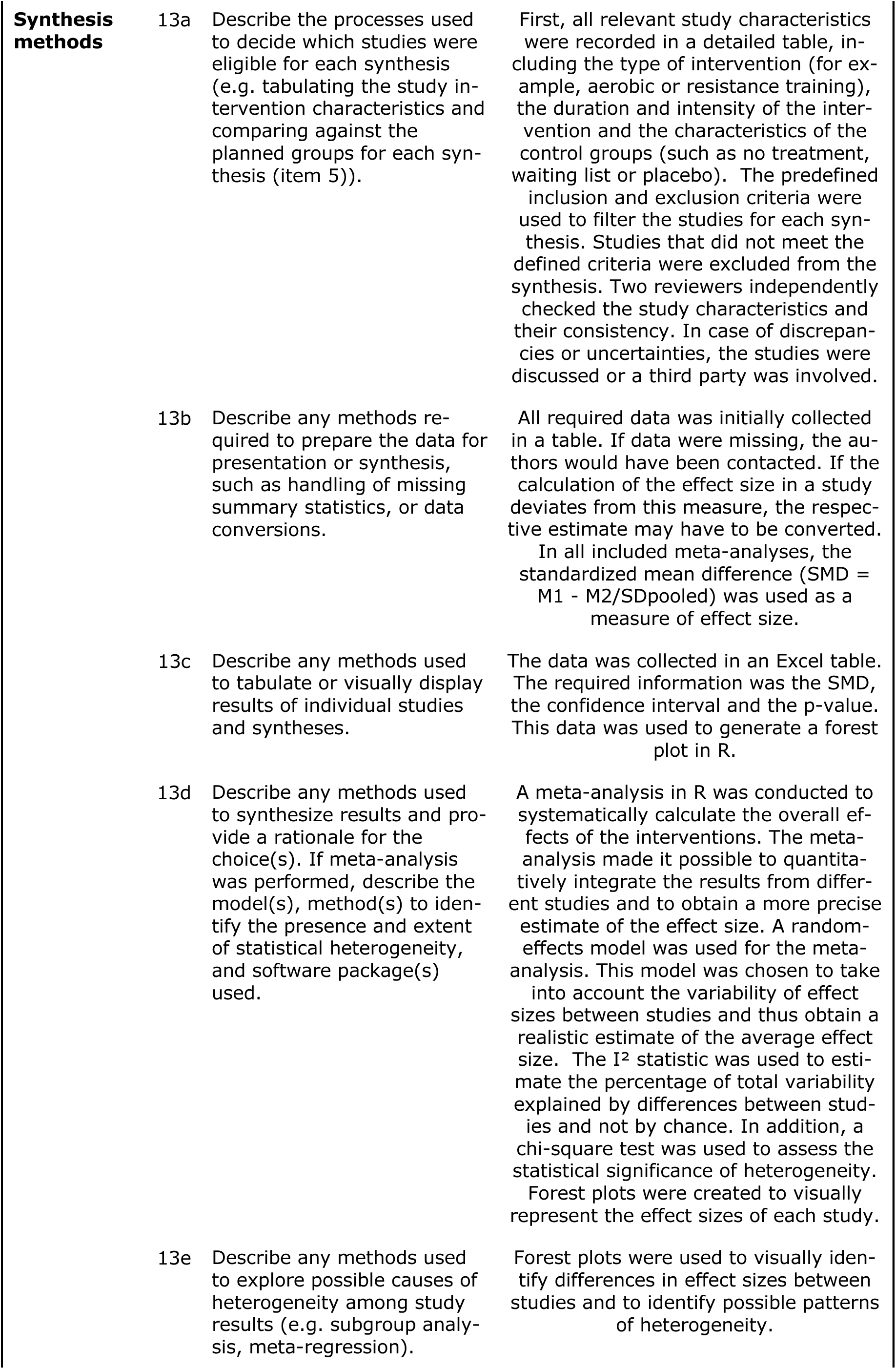
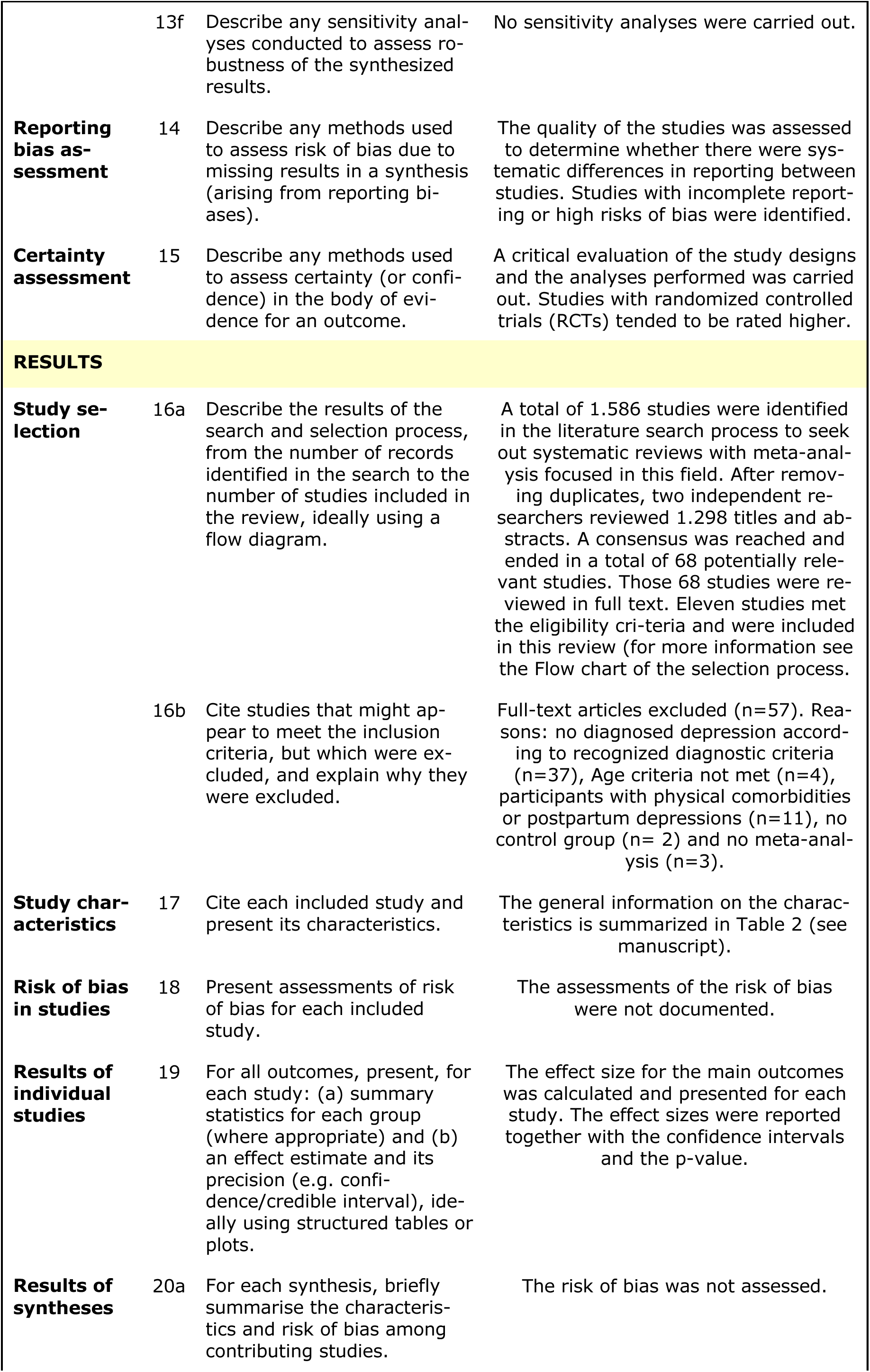
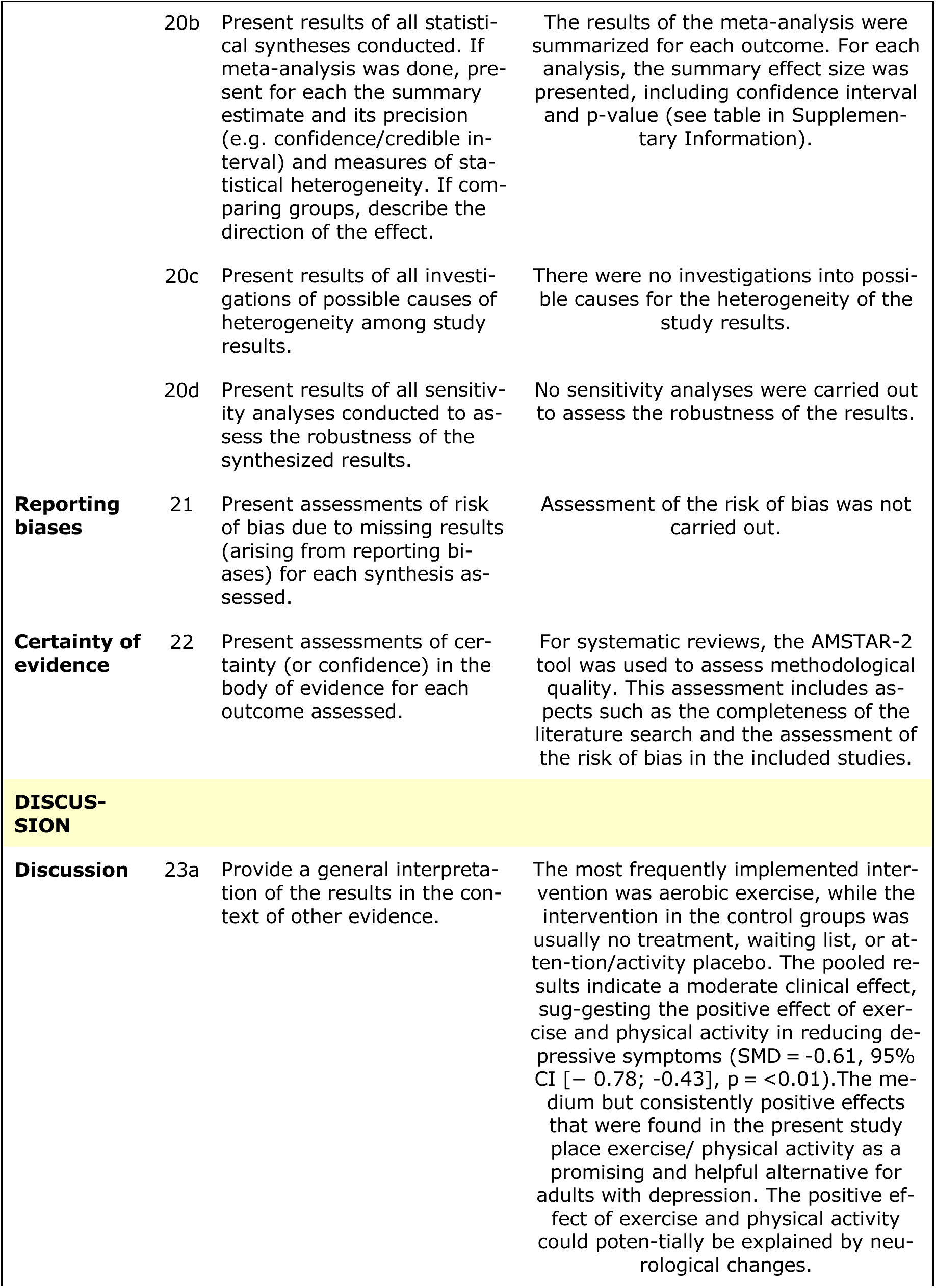
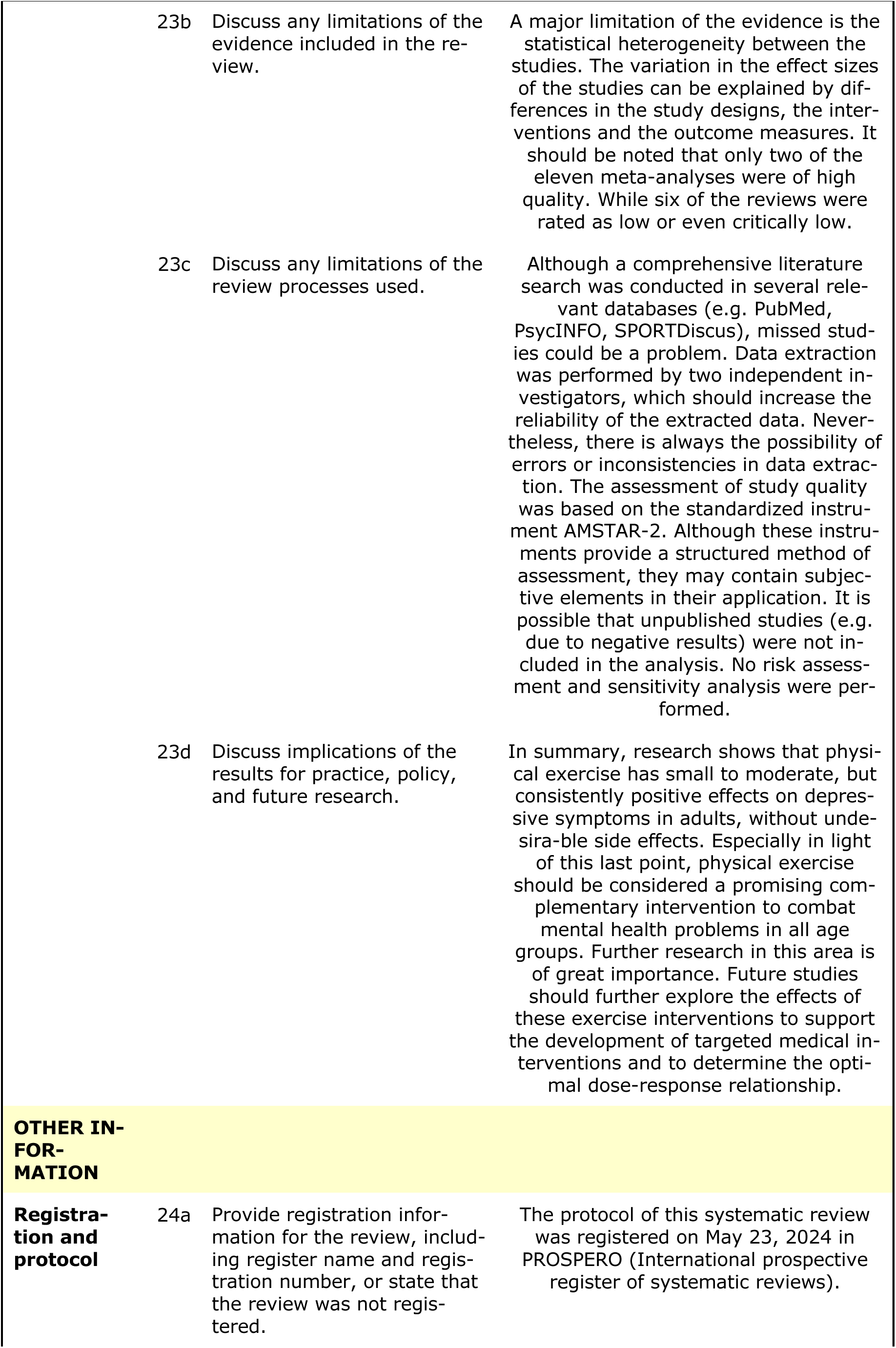
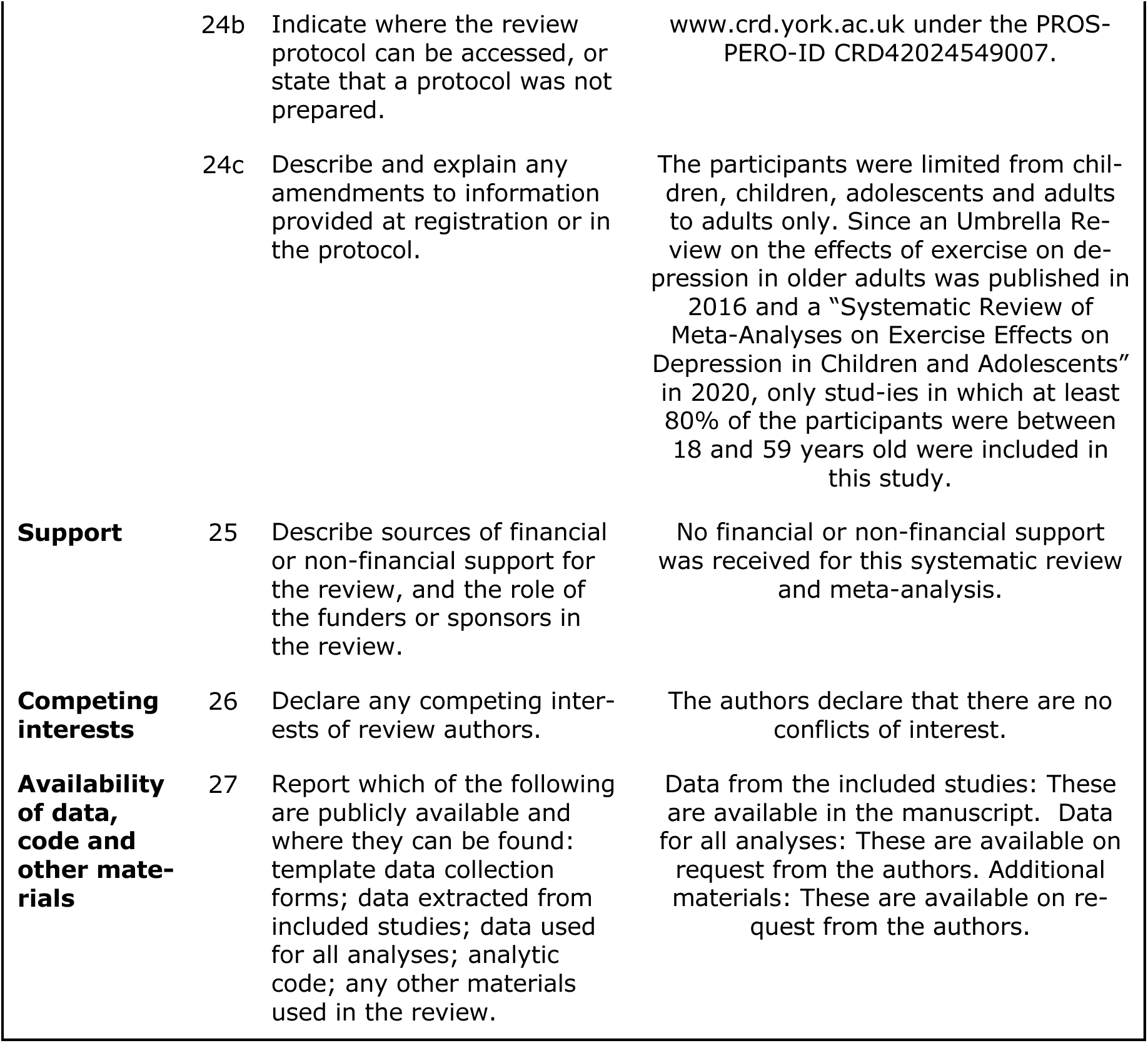
PRISMA 2020 Main Checklist.

**Supplemental 2:** Data for the Forest plot.

| Sutdy | Std. Mean Difference | Lower_CI | Upper_CI | p |
| --- | --- | --- | --- | --- |
| Bailey et al. [27] | -0.82 | -1.02 | -0.61 | 0.050 |
| Correia et al. [5] | -0.76 | -1.18 | -0.34 | 0.010 |
| Guo et al. [47] | -1.13 | -1.48 | -0.78 | 0.010 |
| Heissel et al. [30] | -0.946 | -1.18 | -0.71 | 0.001 |
| Jazayeri et al. [23] (Resistance) | -0.14 | -0.16 | -0.12 | 0.000 |
| Jazayeri et al. [23] (Endurance) | -0.16 | -0.18 | -0.14 | 0.000 |
| Ju et al. [45] | -0.71 | -0.90 | -0.52 | 0.020 |
| Krogh et al. [43] | -0.66 | -0.86 | -0.46 | 0.001 |
| Lee et al. [46] | -0.62 | -0.86 | -0.37 | 0.000 |
| Morres et al. [8] | -0.79 | -1.01 | -0.57 | 0.000 |
| Pérez-López et al. [44] | -0.36 | -0.52 | -0.21 | 0.002 |
| Recchia et al. [14] | -0.45 | -0.67 | -0.23 | 0.003 |

